# Early immune activation in the prediagnostic phases of immune-mediated neurological diseases

**DOI:** 10.64898/2026.06.26.26356707

**Authors:** Hannes Vietzen, Raphael Reinecke, Jan P. Nolte, Laura M. Kushner, Sarah M. Berger, Jeremy V. Camp, Markus Ponleitner, Kevin Rostásy, Henrieke Saucke, Franziska Kauth, Georgia Koukou, Simon Sommer, Eva-Maria Wendel, Marianne Graninger, Verena Endmayr, Katrin Koebl-Shkreli, Sophie Nitsch, Johanna Wachutka, Emmanuelle L. Waubant, Soe Mar, Amy T. Waldman, Lauren B. Krupp, T. Charles Casper, Teri Schreiner, Tanuja Chitnis, Lisa Weidner, Charlotte Pistorius, Christof Jungbauer, Markus Reindl, Barbara Kornek, Markus Breu, Gabriel Bsteh, Hans Lassmann, Elisabeth Puchhammer-Stöckl, Thomas Berger, Romana Höftberger, Paulus Rommer

**Author notes:** **Corresponding Author: Hannes Vietzen**, **PhD**, Center for Virology, Medical University of Vienna, Kinderspitalgasse 15, 1090 Vienna, Austria.

## Abstract

Multiple sclerosis (MS), myelin oligodendrocyte glycoprotein antibody–associated disease (MOGAD), and neuromyelitis optica spectrum disorder (NMOSD) are immune-mediated inflammatory disorders of the central nervous system (CNS). The temporal relationship between disease-specific autoantibodies and biomarkers of CNS injury before diagnosis remains unclear and is relevant for understanding early pathobiology. Here, we conducted a multicentre retrospective longitudinal case-control study using prediagnostic plasma from 362 individuals who later developed MS, 145 who developed MOGAD, and 60 who developed NMOSD.

Plasma IgG levels against CNS antigens, MOG, and AQP4, as well as neurofilament light chain (pNfL), were quantified, and temporal relationships between immune activation, neuroaxonal injury, and clinical disease onset were modelled using linear mixed-effects models and survival analyses.

In MS, EBNA-1–specific and CNS-cross-reactive IgG were elevated up to 77.8 months before diagnosis, preceding pNfL increases by 44.9 months. In NMOSD, AQP4-IgG seroconversion occurred 32.5 months before diagnosis and preceded pNfL elevations by 40.4 months. In MOGAD, pNfL elevations preceded MOG-IgG seroconversion by 11.2 months.

Thus, in MS and NMOSD, humoral autoimmunity precedes detectable CNS injury, whereas in MOGAD, neuroaxonal injury occurs before circulating MOG-IgG. These distinct temporal patterns suggest differing early immunopathological trajectories and may provide a framework for future studies of early disease biology and biomarker-guided risk stratification.

## Introduction

Multiple sclerosis (MS), neuromyelitis optica spectrum disorder (NMOSD), and myelin oligodendrocyte glycoprotein antibody-associated disease (MOGAD) are immune-mediated inflammatory disorders of the central nervous system (CNS). Although they share overlapping clinical and imaging features, they differ markedly in immunopathogenesis and treatment response. The identification of aquaporin-4-specific immunoglobulin G (AQP4-IgG) has refined diagnostic classification and mechanistic concepts for NMOSD, whereas myelin oligodendrocyte glycoprotein-specific IgG (MOG-IgG) serves as the defining serological marker of MOGAD ^1–6^.

Epstein-Barr virus (EBV), a ubiquitous herpesvirus, has been consistently associated with the risk of developing MS. Large longitudinal studies have demonstrated that EBV seroconversion precedes MS diagnosis by several years, with reported median intervals ranging from 6.4 to 8.2 years ^7–9^. Immune responses to the EBV nuclear antigen 1 (EBNA-1), particularly to the conserved EBNA-1_381–452_ peptide, have been implicated as markers of altered immune tolerance in individuals who later develop MS. A longitudinal study using archived serial prediagnostic samples showed that persistently increased IgG responses against EBNA-1_381-452_ occurred exclusively in individuals who later developed MS ^8^. Antibodies directed against EBNA-1_381-452_ can cross-react with several CNS-associated antigens, including glial cell adhesion molecule (GlialCAM), α-crystallin B chain (CRYAB), myelin basic protein (MBP), and anoctamin 2 (ANO2) _8,10-14._

Despite advances in antibody detection, the temporal relationships between antibody emergence and markers of CNS injury before diagnosis remain poorly defined in NMOSD and MOGAD. Prediagnostic AQP4-IgG positivity has been reported, but evidence is largely restricted to isolated case reports or small case series ^15,16^. Systematic longitudinal data on prediagnostic neurofilament light chain (NfL) or glial fibrillary acidic protein (GFAP) concentrations, biomarkers of axonal damage and astrocytic injury, are lacking for NMOSD and MOGAD.

In this study, we analysed prediagnostic longitudinal plasma samples from individuals who later developed MS, NMOSD, or MOGAD to define the temporal relationships between early antibody responses, blood biomarkers of CNS injury, and clinical diagnosis between early immune responses and downstream biological changes across these immune-mediated CNS disorders.

## Material and Methods

### Study Participants

This multicentre, retrospective, longitudinal matched case–control study included patients diagnosed with immune-mediated inflammatory disorders of the central nervous system (CNS) between 2001 and 2023. The study was based on archived plasma samples obtained before and after clinical MS, NMOSD, or MOGAD diagnosis.

Patients were identified from established clinical cohorts and institutional registries at the Medical University of Vienna (N=819, 60.8%), the BIOMARKER study at the Children’s Hospital Datteln, Germany (N=338, 25.1%), and the US Network of Paediatric MS Centres (N=190, 14.1%), yielding a total cohort of 1,347 individuals. MS was diagnosed according to the 2017 McDonald criteria ^17,18^. NMOSD was diagnosed according to the 2015 international consensus diagnostic criteria,^19^ and all included NMOSD patients tested positive for aquaporin-4 immunoglobulin G (AQP4-IgG) as determined by a live cell-based assay (LCBA) transfected with the M23 isoform. MOGAD was defined by detection of MOG-specific IgG using an LCBA transfected with full-length native MOG, in accordance with proposed diagnostic criteria ^4^. MOG-IgG positivity was defined by a high-titre cut-off of ≥1:160. Clear positive titres (≥1:640) required fulfilment of core clinical criteria alone, while low-positive titres (1:160-1:320) necessitated the presence of both core and supporting clinical and imaging features ^4^. All LCBAs were performed at certified reference laboratories at the Medical University of Innsbruck and the Medical University of Vienna.

Two sample types were analysed: (i) diagnostic index samples collected at or shortly after diagnosis (or a matched index date in controls) and (ii) archived prediagnostic plasma samples collected before diagnosis (or a matched index date in controls).

Diagnostic index samples from cohorts in Austria, Germany, and the USA were used for cross-sectional comparisons at diagnosis. The diagnostic cohorts comprised 1,039 patients with relapsing–remitting MS (MS), 104 with aquaporin-4 immunoglobulin G–positive NMOSD, and 204 with MOGAD.

Clinical onset was defined as the first documented neurological symptoms attributable to MS, NMOSD, or MOGAD based on the medical record. Diagnosis date was defined as the date on which the respective diagnostic criteria were first fulfilled and recorded.

Archived prediagnostic plasma samples collected before diagnosis (or a matched index date in controls) were available for a subset of patients in the diagnostic cohort (MS: 34.8%; NMOSD: 57.7%; MOGAD: 72.1%). All archived samples had been collected before documented clinical onset.

At least one archived prediagnostic plasma sample was available from 362 MS patients, 60 NMOSD patients, and 145 MOGAD patients, collected at 1.6-75.6 (median 22), 0.03-35.8 (median 17), and 0.6-37 (median 8.4) months before diagnosis, respectively. Across diseases, the interval between clinical onset and diagnosis ranged from 0.1 to 22.7 months (median 1.9). On average, 3.1 (range 1-4), 1.7 (range 1-4), and 1.7 (range 1-4) plasma samples per patient were available for MS, NMOSD, and MOGAD, respectively, within the 10 years preceding clinical diagnosis. All archived prediagnostic plasma samples were collected before recorded clinical disease onset and disease diagnosis. Prediagnostic plasma samples originated from routine clinical care and were subsequently retrieved from institutional biobanks; they were not collected specifically for this study. Archived prediagnostic patient samples were obtained from institutional biobanks and blood service repositories in Austria. Most prediagnostic patient samples were originally collected for routine virological diagnostics—such as screening for viral infections and serological analyses, including vaccination titer controls—and were subsequently stored at the Biobank of the Center for Virology, Medical University of Vienna.

Total EBNA-1-specific IgG as well as pNfL and pGFAP measurements from prediagnostic plasma samples from 39 patients with MS (10.8%) have been reported previously ^7,8^. All archived prediagnostic plasma samples from NMOSD and MOGAD patients were tested for the presence of AQP4-IgG and MOG-IgG. Positive samples were subsequently titrated.

Each patient was matched 1:1 to a healthy control by age and sex, and by availability of archived plasma within a predefined sampling window relative to the patient’s index date. All healthy controls were selected from the Austrian blood service repositories and the biobank of the Center for Virology, Medical University of Vienna. Healthy controls were selected from individuals without known neuroinflammatory disease at the time of sampling. Available longitudinal follow-up information confirmed that none of the control individuals developed a neuroinflammatory disorder during the observation period. For MS, matching additionally accounted for EBV serostatus and timing of EBV seroconversion. Details of the included sampling time points are shown in Table S1, characteristics of this study cohort are provided in Table S2.

### Serology

EBV-EBNA-1-IgG and EBV-VCA-IgG were detected and quantified by enzyme-linked immunosorbent assay (ELISA, Euroimmun and DRG International). Pooled GlialCAM_370-389_-, CRYAB_2-21_-, MBP_205-224_-, and ANO2_135-154_-specific IgG (referred to as CNS-specific IgG, Table S3) were detected and quantified by ELISA as described previously.^8^ IgG titres were assessed in two independent technical replicates.

### NfL and GFAP Testing

PNfL and pGFAP were quantified using single-molecule array (SIMOA) Neurology 2 Plex B assay kits in the SIMOA SR-X Analyzer (Quanterix) according to the manufacturer’s instructions.^20^

## Data Availability Statement

De-identified data are available from the corresponding author upon request and upon approval by the data-clearing committee of the Medical University of Vienna.

## Ethics Declaration

The study was approved by the local Institutional Review Boards (IRB numbers: 1123/2015, 1133/2022 1339/2022, 1668/2023, 164/2014, AN4059, #10-05039).

## Statistical Analysis

The study size was determined by the availability of retrospectively collected prediagnostic plasma samples from individuals who later developed MS, NMOSD, or MOGAD and from matched healthy controls. EBV IgG seroprevalence was compared using Fisher’s exact test. Cross-sectional group comparisons were performed using unpaired t-tests with Welch’s correction, where appropriate. Within-individual changes across repeated prediagnostic time points were analysed using non-parametric tests for repeated measures (Friedman test with Dunn’s post hoc correction).

Antibody seroprevalence over time was analysed using Kaplan-Meier methods. Antibody and biomarker trajectories over time were analysed using linear mixed-effects models including fixed effects for group, time (modelled using cubic splines), and their interaction, with a random intercept to account for repeated measurements within individuals. Pairwise comparisons of estimated marginal means at predefined three-month time points were adjusted for multiple testing using the Šídák correction.

All available samples with sufficient volume and valid assay performance (i.e., successful measurement without technical failure) were included in the analyses. No additional exclusions were applied. All variables included in the statistical analyses had complete data. Statistical analyses were performed using GraphPad Prism 10 or SPSS Statistics 27. Linear mixed-effects models were fitted using the lme4 (v1.1.37) and emmeans (v1.11.1) packages in R (v4.5.2). Two-sided p values < 0.05 were considered statistically significant.

## Results

### Temporal Dynamics of Antibodies before MS, NMOSD, and MOGAD Diagnosis

We first assessed the temporal emergence of disease-relevant antibodies in plasma samples collected within 10 years before clinical diagnosis of MS, NMOSD, or MOGAD (Fig.1A). These samples were tested for EBNA-1-specific IgG, AQP4-IgG, and MOG-IgG, respectively. As demonstrated by Kaplan–Meier analyses, EBNA-1-specific IgG in individuals who later developed MS reached seroprevalence above 50% at a median of 77.8 months before diagnosis (Fig.1B). In NMOSD, more than 50% of patients with available prediagnostic samples were AQP4-IgG seropositive at a median of 32.5 months before diagnosis (Fig.1C). In contrast, MOG-IgG became detectable only shortly before diagnosis, with patient-level seroprevalence exceeding 50% 7 months before MOGAD diagnosis (Fig.1D).

**Figure 1:**
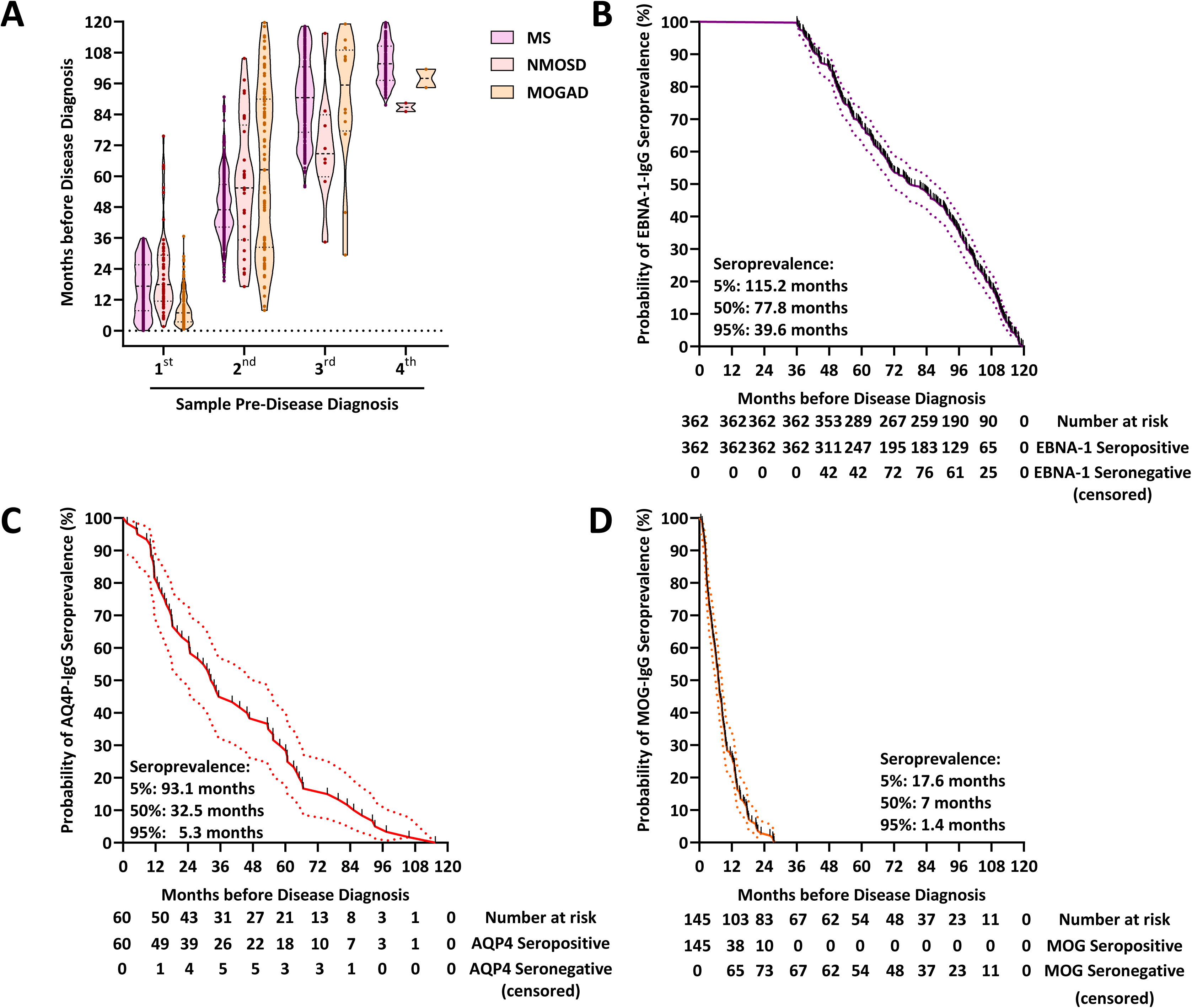
Temporal dynamics of antibody emergence in the preclinical phases of MS, NMOSD, and MOGAD. (A) Retrospective plasma samples were collected from 362 MS patients (mean 3.1, range 1–4 samples per patient), 60 NMOSD patients (mean 1.7, range 1–4), and 145 MOGAD patients (mean 1.7, range 1–4) within ten years before diagnosis. Violin plots depict the distribution of sampling time points (months before diagnosis). Each dot represents an individual plasma sample from an individual future MS, NMOSD, or MOGAD patient. All prediagnostic samples were collected before clinical onset. (B–D) All plasma samples from future (B) MS, (C) NMOSD, and (D) MOGAD patients were tested for the presence of EBNA-1-specific–IgG, AQP4-IgG, and MOG-IgG, respectively. Seroprevalence over time was analysed using Kaplan-Meier curves. Kaplan–Meier analyses include only individuals with at least one prediagnostic plasma sample (MS n=362; NMOSD n=60; MOGAD n=145). **Abbreviations**: **AQP4:** Aquaporin-4, **EBNA-1:** Epstein-Barr Virus Nuclear Antigen 1, **MOG:** Myelin Oligodendrocyte Glycoprotein, **MOGAD:** Myelin Oligodendrocyte Glycoprotein Antibody–Associated Disease, **MS**: Multiple Sclerosis, **NMOSD**: Neuromyelitis Optica Spectrum Disorder.

### Temporal Dynamics of pNfL and pGFAP before MS, NMOSD, and MOGAD Diagnosis

We next examined longitudinal trajectories of pNfL and pGFAP relative to diagnosis. Each patient was matched 1:1 to a healthy control according to age, sex, and the availability of archived plasma samples collected before the patient-specific index date (Fig.2A-C). pNfL and pGFAP were measured in all available plasma samples obtained before (first to fourth sample) and after diagnosis (Fig.S1).

**Figure 2:**
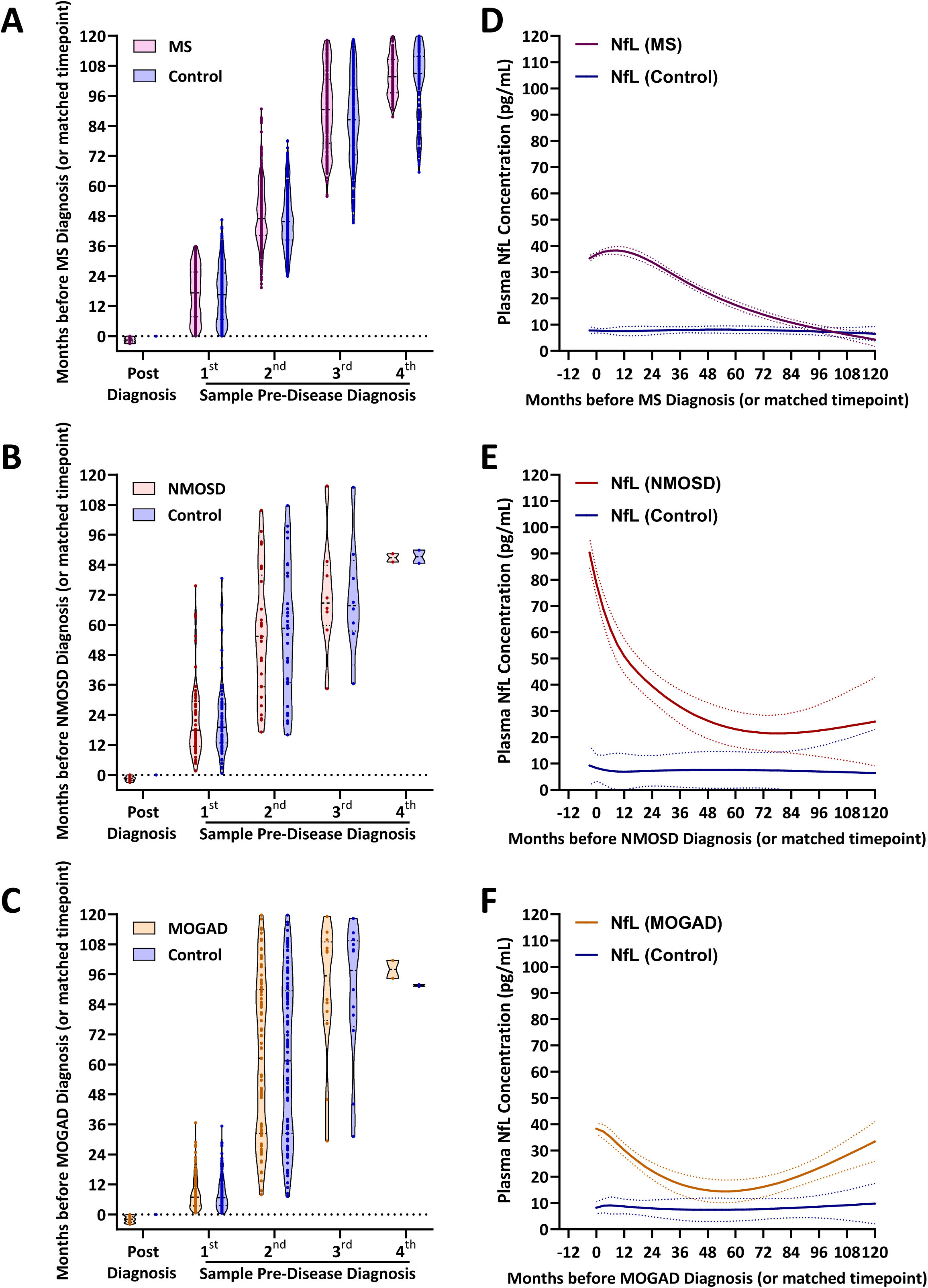
Temporal dynamics of pNfL in the preclinical phases of MS, NMOSD, and MOGAD. Plasma samples were collected from (A) 1,039 MS patients and matched controls, (B) 104 NMOSD patients and matched controls, and (C) 204 MOGAD patients and matched controls after the respective disease diagnoses. In addition, retrospective plasma samples were collected from (A) 362 MS patients and matched controls (mean 3.1, range 1–4 samples per patient), (B) 60 NMOSD patients and matched controls (mean 1.7, range 1–4), and (C) 145 MOGAD patients and matched controls (mean 1.7, range 1–4) within ten years before diagnosis or a matched time point for controls. Violin plots depict the distribution of sampling time points (months before and after diagnosis). Each dot represents an individual plasma sample from a future MS, NMOSD, or MOGAD patient, or from a matched control. (D–F) All plasma samples from (D) MS and matched controls, (E) NMOSD and matched controls, and (F) MOGAD patients and matched controls were tested for pNfL concentrations. pNfL concentrations were analysed using a linear mixed model to account for repeated sampling. Means and standard error are shown. **Abbreviations**: **MOGAD:** Myelin Oligodendrocyte Glycoprotein Antibody–Associated Disease, **MS**: Multiple Sclerosis, **NfL**: neurofilament light chain, **NMOSD**: Neuromyelitis Optica Spectrum Disorder.

As shown in Fig.S2, pNfL and pGFAP concentrations were significantly higher after diagnosis in all three disorders compared with matched controls. Notably, elevations were also observed before diagnosis, with patients exhibiting higher pNfL and pGFAP levels than controls during the prediagnostic period.

As shown in Fig.2D-F by longitudinal mixed-effects models, pNfL concentrations in future MS, NMOSD, and MOGAD patients exceeded control levels from 69, 27, and 21 months before diagnosis, respectively. pGFAP concentrations exceeded control levels from 75 months before diagnosis in MS and from 45 months before diagnosis in MOGAD, whereas in NMOSD, increased pGFAP concentrations were observed only from 6 months before diagnosis (Fig.S3; all Šídák-adjusted p<0.04).

### Temporal Relationship between Antibody Emergence and Increase of pNfL and pGFAP before MS, NMOSD, and MOGAD Diagnosi

We then assessed the temporal relationship between the emergence of disease-specific antibodies and subsequent elevations in pNfL and pGFAP. High pNfL and pGFAP levels were defined as concentrations exceeding the 95^th^ percentile of healthy controls (pNfL ≥14.91 pg/mL; pGFAP ≥121 pg/mL, Fig.S4A-B).

For each participant, the interval between first antibody detection and first biomarker elevation was then calculated. In MS and NMOSD, the appearance of EBNA-1-specific IgG and AQP4-IgG preceded elevations in pNfL and pGFAP by mean intervals of 44.9 and 47.3 months, and 40.4 and 38.7 months, respectively (Fig.S4C-D). By contrast, in individuals who later developed MOGAD, elevated pNfL and pGFAP levels were observed a mean of 11.2 and 9.8 months before the emergence of MOG-IgG.

### CNS-specific IgG Titres before MS Diagnosis

We next evaluated longitudinal trajectories of EBNA-1-derived CNS-specific IgG titres in MS. CNS-specific IgG titres were quantified in all available plasma samples (Fig.S5A). Across all prediagnostic time points, CNS-specific IgG titres were significantly higher in future MS patients than in matched controls (Fig.3A and Fig.S5B; all Šídák-adjusted p<0.0006) and increased further as the time of MS diagnosis approached.

**Figure 3:**
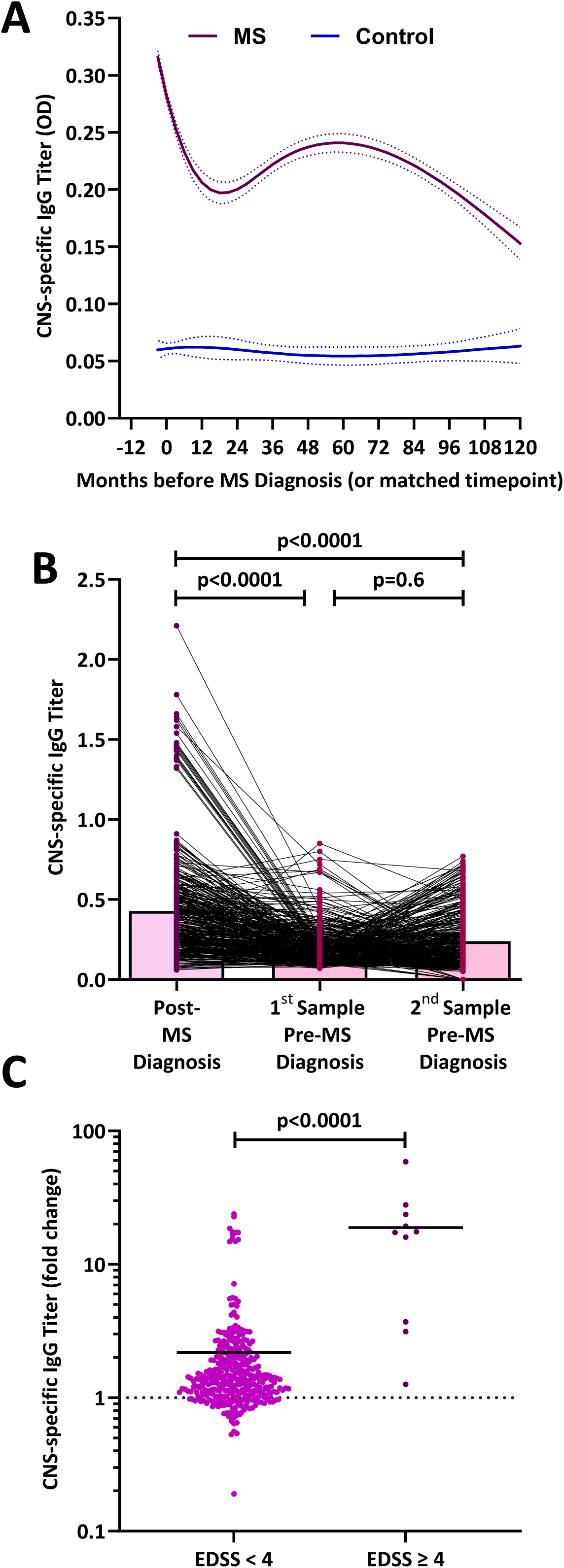
Temporal dynamics of CNS-specific IgG antibodies in the preclinical phase of MS. Plasma samples were collected from 1,039 MS patients after diagnosis and from matched controls. In addition, retrospective plasma samples were collected from 362 MS patients and matched controls (mean 3.1, range 1–4 samples per patient) within ten years before diagnosis or a matched time point for controls. All plasma samples were tested for CNS-specific IgG titre. (A) CNS-specific IgG titres were analysed using a linear mixed model with a random intercept to account for repeated sampling. Means ± standard error are shown. (B) CNS-specific IgG titres were assessed in 362 MS patients with at least two plasma samples available before diagnosis. Box plots represent the mean CNS-specific IgG titer at each time point, and each dot represents an individual plasma sample. Individual titres over time were compared using the paired Friedman test with Dunn’s multiple comparisons test. (C) CNS-specific IgG titres were assessed in the first plasma sample before and after MS diagnosis. Data are shown as fold change of CNS-specific IgG titres after vs. before diagnosis, stratified by patients with EDSS ≥4 or EDSS <4. Each dot represents an individual plasma sample. Titres were compared using the Mann-Whitney test. **Abbreviations**: **CNS:** central nervous system, **EDSS:** expanded disability status scale**, MS**: Multiple Sclerosis.

To examine within-individual changes, CNS-specific IgG titres obtained before and after diagnosis were compared. Titres were significantly higher after diagnosis, whereas no consistent increase was observed across the two assessed prediagnostic time points (Fig.3B). When stratified by disability at diagnosis, patients with EDSS ≥4 showed a greater prediagnostic increase in CNS-specific IgG titres than patients with EDSS <4 (Fig.3C).

### Temporal Dynamics of CNS-IgG Titres before NMOSD and MOGAD Diagnosis

We subsequently assessed CNS-specific IgG trajectories in NMOSD and MOGAD. At diagnosis, CNS-specific IgG titres were significantly elevated in both disorders (Fig.S6A–D). During the prediagnostic period, titres exceeded control levels from 27 months before diagnosis in NMOSD and from 3 months before diagnosis in MOGAD (Fig.S7E–F; Šídák-adjusted p<0.04). In both disorders, these increases occurred after the initial detection of AQP4-IgG or MOG-IgG, respectively.

### Temporal Dynamics of AQP4-IgG in NMOSD Patients

We next examined longitudinal changes in AQP4-IgG titres in NMOSD patients. Therefore, all AQP4-IgG-positive plasma samples were quantified using an LCBA (Fig.S7A). Mean titres were significantly higher after diagnosis than at all prediagnostic time points (Fig.S8B). ROC analysis did not identify an absolute titre threshold distinguishing prediagnostic from clinical NMOSD (Fig.S8C).

We therefore assessed relative within-individual changes. All NMOSD patients exhibited a rise in AQP4-IgG titres immediately before diagnosis (Fig.4A). Patients with EDSS ≥4 showed a greater prediagnostic titre increase than those with EDSS <4 (Fig.4B).

**Figure 4:**
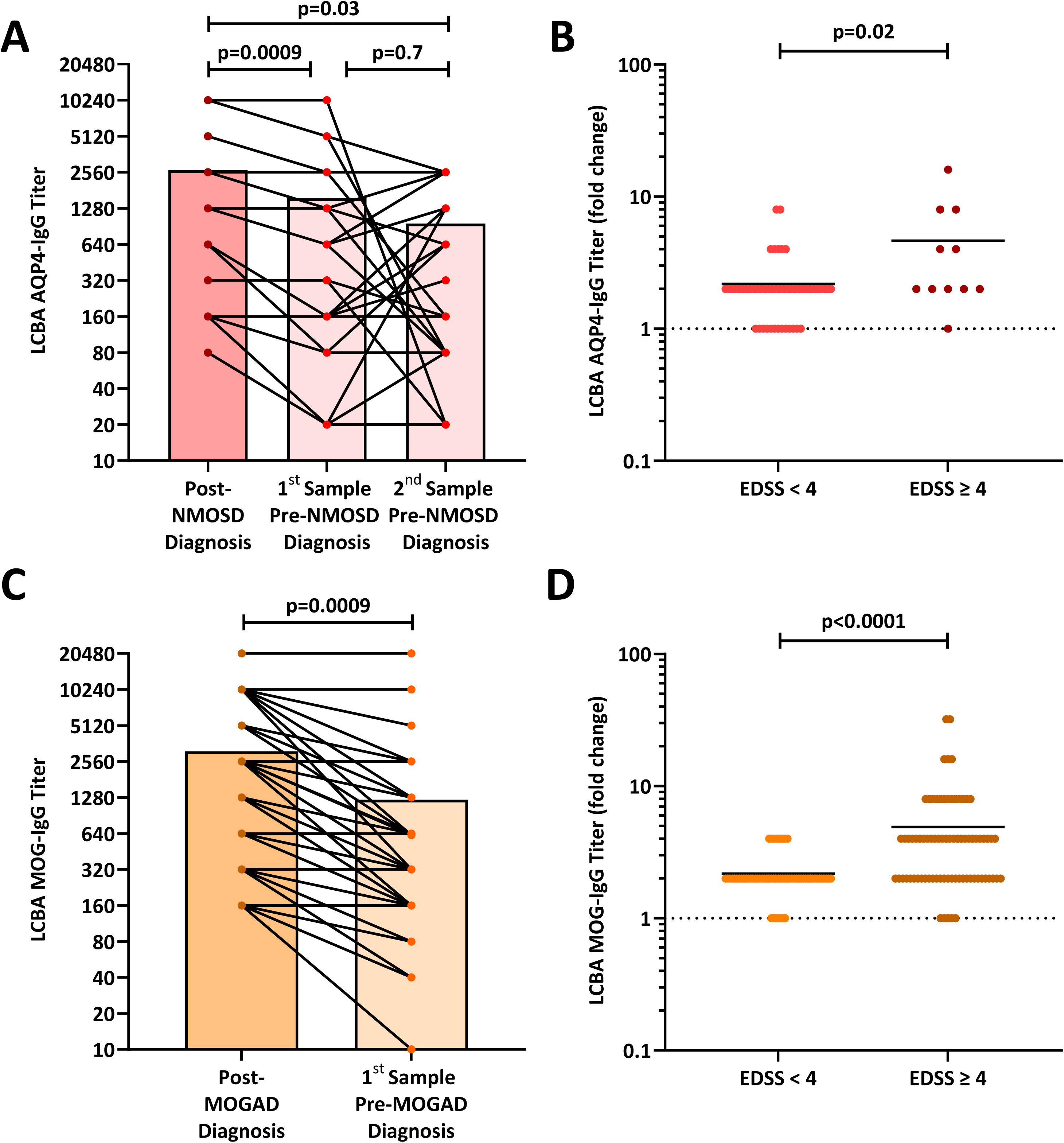
Temporal dynamics of AQP4-and MOG-IgG in the preclinical phases of NMOSD and MOGAD. (A) AQP4-specific IgG titres were assessed in 29 NMOSD patients with at least two plasma samples available before diagnosis. Box plots represent the mean AQP4-specific IgG titer at each time point, and each dot represents an individual plasma sample. Individual titres over time were compared using the paired Friedman test with Dunn’s multiple comparisons test. (B) AQP4-specific IgG titres were assessed in the first plasma sample before and after NMOSD diagnosis. Data are shown as fold change of AQP4-specific IgG titres after vs. before diagnosis, stratified by patients with EDSS ≥4 or EDSS <4. Each dot represents an individual plasma sample. Titres were compared using the Mann-Whitney test. (C) MOG-specific IgG titres were assessed in 141 MOGAD patients with at least two plasma samples available before diagnosis. Box plots represent the mean MOG-specific IgG titer at each time point, and each dot represents an individual plasma sample. Individual titres over time were compared using the paired Friedman test with Dunn’s multiple comparisons test. (D) MOG-specific IgG titres were assessed in the first plasma sample before and after MOGAD diagnosis. Data are shown as fold change of MOG-specific IgG titres after vs. before diagnosis, stratified by patients with EDSS ≥4 or EDSS <4. Each dot represents an individual plasma sample. Titres were compared using the Mann-Whitney test. **Abbreviations**: **AQP4:** Aquaporin-4**, EDSS:** Expanded Disability Status Scale**, LCBA:** live cell-based assay**, MOG:** Myelin Oligodendrocyte Glycoprotein, **MOGAD:** Myelin Oligodendrocyte Glycoprotein Antibody–Associated Disease**, NMOSD:** Neuromyelitis Optica Spectrum Disorder.

### Temporal Dynamics of MOG-IgG Titres before MOGAD Diagnosis

Finally, we analysed MOG-IgG trajectories in MOGAD. Therefore, all MOG-IgG-positive plasma samples were quantified using an LCBA (Fig.S8A). Mean titres were higher after diagnosis than at the prediagnostic time point (Fig.S8B). An increase in MOG-IgG titres before diagnosis was observed in 91.4% of patients (Fig.4C). Prediagnostic titre increases were greater in patients with EDSS ≥4 than in those with EDSS <4 (Fig.4D). Together, these findings indicate that while MOG-IgG emergence does not precede early biomarker evidence of neuroaxonal injury, relative increases in MOG-IgG titres are associated with progression to clinically manifest MOGAD.

## Discussion

This study provides one of the most comprehensive analyses to date of prediagnostic immune activation in MS, NMOSD, and MOGAD, by integrating longitudinal antibody profiling with blood biomarkers of neuroaxonal and astrocytic injury. Using serial prediagnostic samples, we describe disease-specific temporal patterns of antibody emergence and their relationship to subsequent increases in NfL and GFAP. These findings extend, and in some aspects refine, current concepts of the earliest immunopathological events in these disorders.

First, our data demonstrate a prolonged prediagnostic phase in MS and NMOSD, characterised by the early emergence of CNS-directed antibody responses, including EBNA-1-related CNS-cross-reactive IgG in MS and AQP4-IgG in NMOSD, occurring years before clinical diagnosis and well before detectable increases in pNfL or pGFAP. EBNA-1-specific IgG responses appeared approximately 6.5 years before MS diagnosis, consistent with previous reports implicating EBNA-1–mediated immune dysregulation in the prediagnostic stages of MS ^7–9,21–23^. The sustained elevation of EBNA-1–derived CNS-specific antibodies observed across the prediagnostic period suggests persistent immune activation or impaired tolerance, mediated by molecular mimicry to CNS-derived antigens.

In contrast to the EBV-associated trajectory observed in MS, the early immunopathogenesis of NMOSD followed a distinct pattern. Our results confirm and extend earlier observations of prediagnostic AQP4-IgG seropositivity ^15,16^. At a cohort level, AQP4-IgG not only emerged years before diagnosis but increased steadily over time. The relative increase, rather than the absolute antibody titre, was associated with imminent clinical presentation and higher disability at diagnosis. This pattern is consistent with astrocytopathic mechanisms involving complement activation and antibody-dependent cellular cytotoxicity (ADCC) ^24^. Experimental data demonstrating CD40-dependent central tolerance to AQP4 provide a plausible biological framework for the gradual prediagnostic expansion of AQP4-reactive immunity ^25^. Peripheral recognition of AQP4 may further amplify systemic antibody responses ^26,27^. Secondary increases in other CNS-specific IgG may reflect humoral immune responses emerging after immune-mediated tissue damage. Second, and in clear contrast to MS and NMOSD, the immunobiology of MOGAD appeared fundamentally distinct. MOG-IgG seroconversion occurred only shortly before diagnosis and was preceded by elevations in pNfL and pGFAP in most individuals. Although MOG-IgG can induce demyelination in experimental models and in patients through complement activation and ADCC,^28^ their involvement in the disease initiation differs from that of AQP4-IgG^29^. Experimental studies indicate that MOG antibody-mediated demyelination requires concomitant encephalitogenic T-cell responses and is less efficient in activating complement than AQP4-IgG ^28,29^. The temporal dissociation observed here is consistent with the possibility that circulating MOG-IgG does not represent the earliest detectable immune event in MOGAD, but instead reflects downstream humoral responses to upstream mechanisms already associated with CNS injury, such as innate immune activation, early T-cell responses, or transient barrier dysfunction.

Third, the detection of elevated pNfL and pGFAP months to years before diagnosis across all three disorders indicates that subclinical CNS injury can precede overt clinical manifestation including in conditions traditionally considered attack-driven, such as NMOSD and MOGAD.

Together, these disease-specific temporal patterns refine current models of early CNS autoimmunity. In MS and NMOSD, prolonged prediagnostic antibody responses precede measurable blood markers of CNS injury, whereas in MOGAD early biomarker elevations occur before detectable disease-specific antibodies. This distinction has important implications for biological stratification and early disease characterisation.

Several limitations warrant consideration. The retrospective design resulted in variable sampling intervals and limited numbers of archived prediagnostic samples per individual. Because pNfL and pGFAP are influenced by age, sex, and systemic factors, absolute biomarker concentrations should be interpreted with caution. Nevertheless, the longitudinal study design and comparisons with age-and sex-matched healthy controls support the interpretation of biomarker trajectories at the group level. As an observational study, temporal associations cannot establish causality. Furthermore, prediagnostic samples were defined relative to the date of diagnosis. Although the exact onset of neurological symptoms cannot always be established retrospectively and unrecognised clinical disease before diagnosis cannot be excluded, the consistent temporal trajectories observed across large patient groups are unlikely to be explained solely by differences in diagnostic timing. In addition,-prediagnostic case–control analyses were restricted to Austria. This design strengthens internal comparability but may limit generalisability to other populations. Although the NMOSD cohort represents the largest currently available prediagnostic dataset, the absolute number of patients remains limited owing to the rarity of the disease. Despite these constraints, our study provides the first comparative framework integrating antibody trajectories with NfL and GFAP across MS, NMOSD, and MOGAD.

These disease-specific temporal programmes extend current concepts of early CNS autoimmunity and have important clinical implications. Integrating disease-specific antibody dynamics with blood-based biomarkers of axonal and astrocytic injury may improve early biological stratification and inform targeted surveillance in individuals at increased risk. While not yet applicable to clinical practice, these trajectories outline how prediagnostic immune activation could support earlier recognition in selected high-risk settings.

In conclusion, MS, NMOSD, and MOGAD follow distinct prediagnostic immunological trajectories. EBNA-1-related and AQP4-IgG responses emerge years before clinical diagnosis and precede blood-based markers of CNS injury, whereas in MOGAD biomarker evidence of injury precedes antibody seroconversion. These findings extend the current conceptual framework of autoimmune neuroinflammation and provide a foundation for future studies of very early disease biology and risk stratification.

## Acknowledgments

We thank Kathrin Schanda (Clinical Department of Neurology, Medical University of Innsbruck, Innsbruck, Austria) for the excellent technical assistance.

## Competing Interests Statement

### T. Berger

has participated in meetings sponsored by and received honoraria (lectures, advisory boards, consultations) from pharmaceutical companies marketing treatments for MS: Almirall, Allergan, Bayer, Biologix, Biogen, Bionorica, BMS, Eisai, Genesis, GSK, Horizon, Janssen, Jazz Pharma, MedDay, Merck, Neuraxpharm, Newbridge, Novartis, Octapharma, Roche, Sandoz, Sanofi, Teva, TG Therapeutics and UCB. His institution has received financial support in the past 12 months by unrestricted research grants (Biogen, Bayer, BMS, Merck, Novartis, Roche, Sanofi, Teva) and for participation in clinical trials in multiple sclerosis sponsored by Alexion, Bayer, Biogen, BMS, Merck, Novartis, Roche, Sanofi, Teva.

### G. Bsteh

has received travel funding from Biogen, Merck, Novartis, Roche, Sanofi, and Teva; has received speaker honoraria from Biogen, BMS, Heidelberg Engineering, Janssen, Lilly, Medwhizz, Merck, Neuraxpharm, Novartis, Roche, Sanofi, Teva and Zeiss; has received honoraria for consulting Adivo Associates, Biogen, BMS, Janssen, Merck, Novartis, Roche, Sanofi and Teva. He has received unrestricted research grants from BMS, Merck and Novartis. He serves on the Executive Committee of the European Committee for Treatment and Research in Multiple Sclerosis (ECTRIMS) and the Board of Directors of the International Multiple Sclerosis Visual System Consortium (IMSVISUAL).

### M. Breu

has received speaker honoraria from Sanofi-Genzyme.

### R. Höftberger

reports speaker honoraria from BMS and UCB. The Medical University of Vienna (Austria; employer of Dr. Höftberger) receives payment for antibody assays and for antibody validation experiments organized by Euroimmun (Lübeck, Germany).

### S. Mar

Participated in Operetta 1 and Operetta II study/Roche. No personal compensations.

### J.P. Nolte

has participated in meetings sponsored by, received speaker honoraria or travel funding from Novartis, Biogen, and Neuraxpharm.

### M. Ponleitner

has received speaker or consulting honoraria from Amicus, Sanofi-Aventis, and Novartis and participated in meetings sponsored by and received travel funding from Amicus, Merck, Novartis, and Sanofi-Genzyme, as well as grants for clinical, research, and exchange fellowships awarded by the European Academy of Neurology (EAN) and the Austrian Society of Neurology.

### P. Rommer

reports speaker and consultancy honoraria from A-med, Almirall, Alexion/AstraZeneca, AMGEN, Amicus, Biogen, Merck, neuraxpharm, Novartis, Roche, Sandoz, Sanofi, has received research grants from Amicus, Biogen, Merck, Roche

### K. Rostásy

is consultant of the Operetta II study/Roche and received honoraria for talks from Roche, Merck, Horizon, Octagam.

### M. Reindl

receives research support from Roche Austria. The Medical University of Innsbruck (Austria; employer of MR) receives payments for antibody assays and for antibody validation experiments organized by Euroimmun (Lübeck, Germany)

The **other authors** declare that they have no conflict of interest.

## Author contributions

Conceptualization: H.V., P.R.; Methodology: H.V.; Validation: H.V., R.R., JP.N, LM.K., SM.B., M.P.; Formal Analysis: H.V., JP.N, JV.C.; Investigation: H.V., R.R., LM.K., SM.B., M.P., V.E., K.KS., S.N., J.W.; Data collection and analysis: H.V., R.R.; Resources: K.R., H.S., F.K., G.K., S.S., EM.W., S.M., AT.W., LB.K., L.W., C.P., C.J., M.R., B.K., M.B.,G.B., E.PS., Writing – Original Draft: H.V, P.R.; Writing - Review & Editing: H.V., EM.W., G.B.; H.L.; E.PS., T.B.; R.H.; P.R.

Shared authorship order was determined through joint discussion between the first authors and the senior (last) author.”

## Abbreviations

ADCC: Antibody-Dependent Cellular Cytotoxicity
ANO2: Anoctamin 2
AQP4: Aquaporin-4
CRYAB: α-Crystallin B Chain
EBNA-1: Epstein–Barr Virus Nuclear Antigen 1
EBV: Epstein–Barr Virus
EBV-VCA: Epstein–Barr Virus Viral Capsid Antigen
EDSS: Expanded Disability Status Scale
ELISA: Enzyme-Linked Immunosorbent Assay
GFAP: Glial Fibrillary Acidic Protein
GlialCAM: Glial Cell Adhesion Molecule
IFN-γ: Interferon Gamma
IgG: Immunoglobulin G
IL-2: Interleukin-2
IRB: Institutional Review Board
LCBA: Live Cell-Based Assay
MBP: Myelin Basic Protein
MOG: Myelin Oligodendrocyte Glycoprotein
MOGAD: Myelin Oligodendrocyte Glycoprotein Antibody-Associated Disease
MS: Multiple Sclerosis
NfL: Neurofilament Light Chain
NMOSD: Neuromyelitis Optica Spectrum Disorder
pGFAP: Plasma Glial Fibrillary Acidic Protein
pNfL: Plasma Neurofilament Light Chain
ROC: Receiver Operating Characteristic
SIMOA: Single Molecule Array

---

**Table 1:**
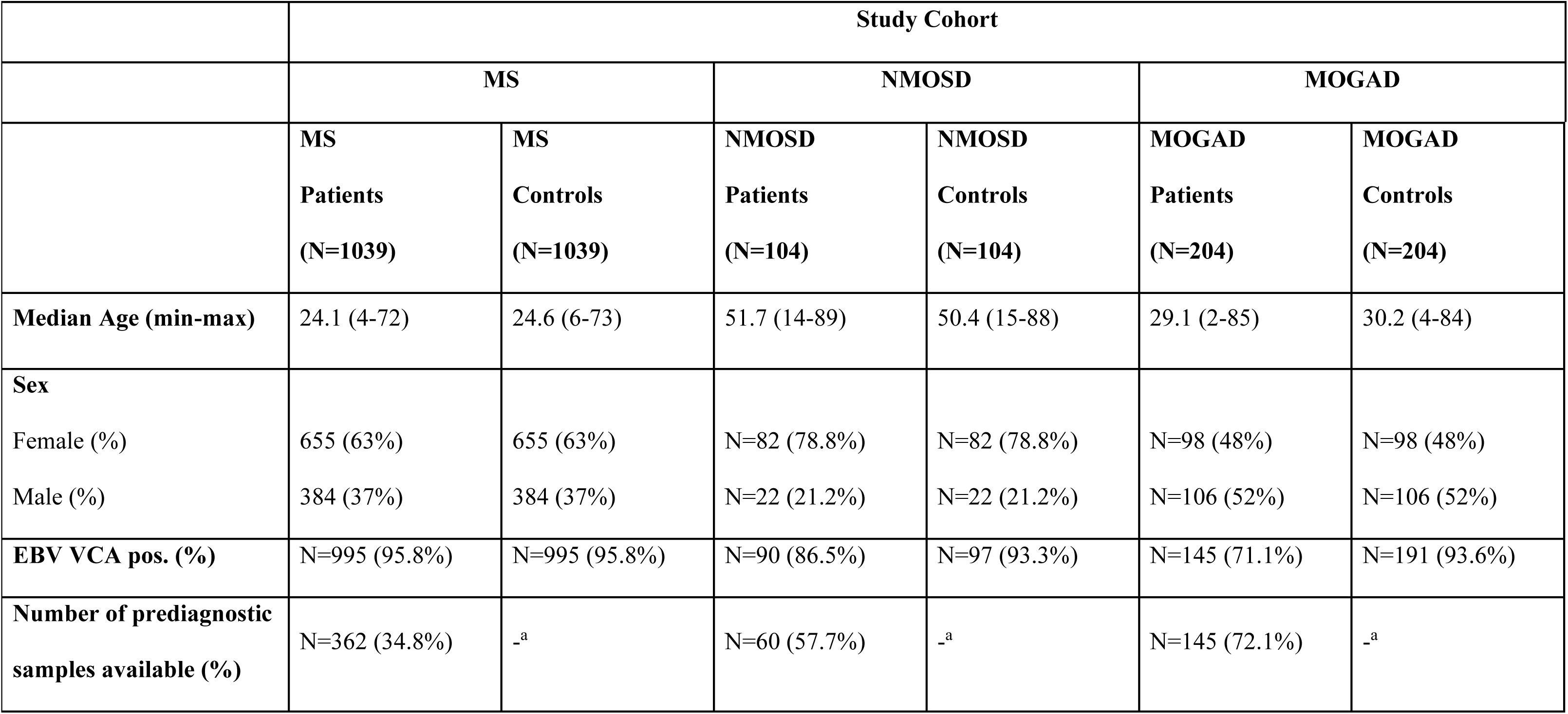
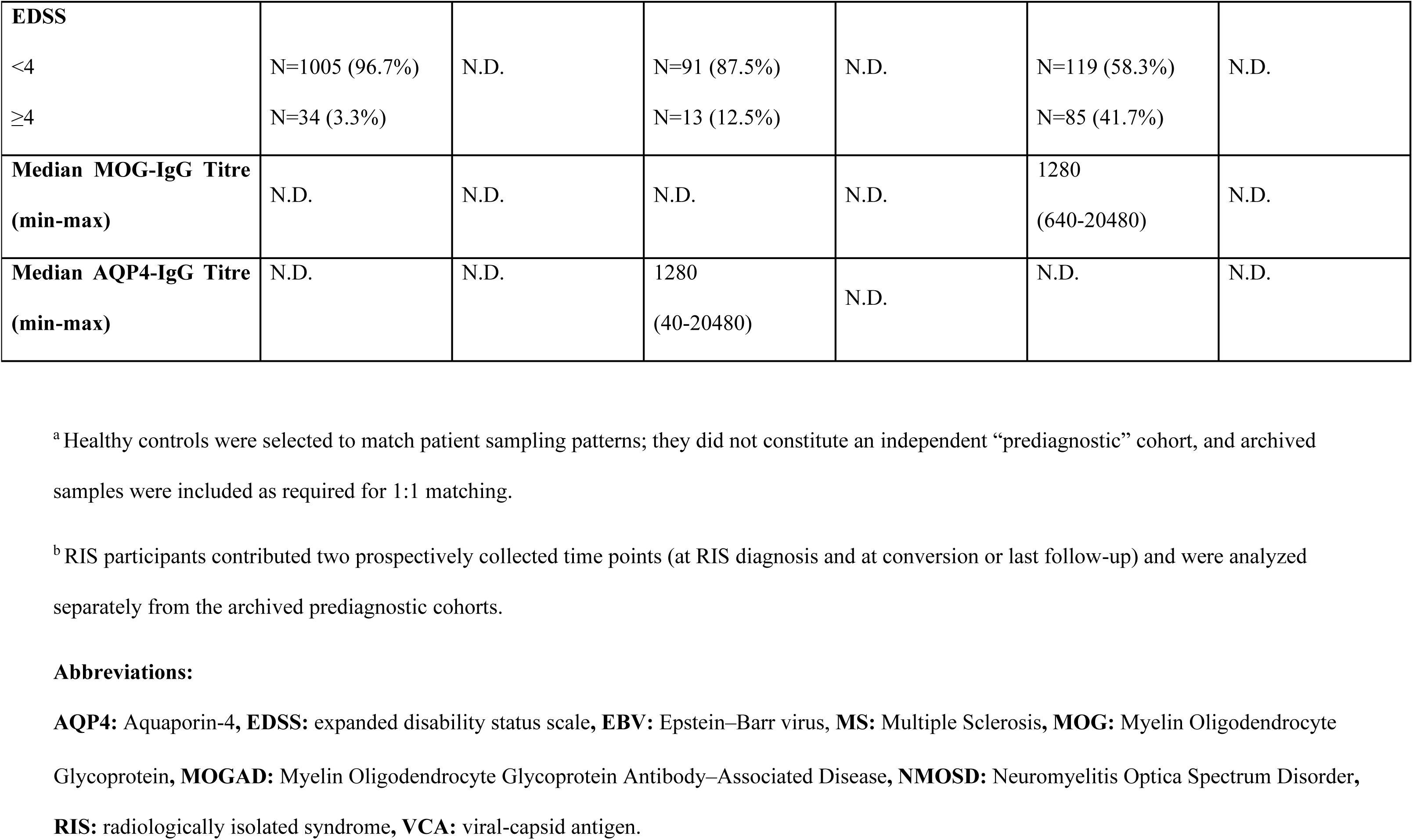
Characteristics of the Study Cohort.

|  | Study Cohort |  |  |  |  |  |
| --- | --- | --- | --- | --- | --- | --- |
|  | MS |  | NMOSD |  | MOGAD |  |
|  | MS<br>Patients<br>(N=1039) | MS<br>Controls<br>(N=1039) | NMOSD<br>Patients<br>(N=104) | NMOSD<br>Controls<br>(N=104) | MOGAD<br>Patients<br>(N=204) | MOGAD<br>Controls<br>(N=204) |
| **Median Age (min-max)** | 24.1 (4-72) | 24.6 (6-73) | 51.7 (14-89) | 50.4 (15-88) | 29.1 (2-85) | 30.2 (4-84) |
| **Sex** |  |  |  |  |  |  |
| Female (%) | 655 (63%) | 655 (63%) | N=82 (78.8%) | N=82 (78.8%) | N=98 (48%) | N=98 (48%) |
| Male (%) | 384 (37%) | 384 (37%) | N=22 (21.2%) | N=22 (21.2%) | N=106 (52%) | N=106 (52%) |
| **EBV VCA pos. (%)** | N=995 (95.8%) | N=995 (95.8%) | N=90 (86.5%) | N=97 (93.3%) | N=145 (71.1%) | N=191 (93.6%) |
| **Number of prediagnostic**<br>**samples available (%)** | N=362 (34.8%) | -<sup>a</sup> | N=60 (57.7%) | -<sup>a</sup> | N=145 (72.1%) | -<sup>a</sup> |
| **EDSS** |  |  |  |  |  |  |
| <4 | N=1005 (96.7%) | N.D. | N=91 (87.5%) | N.D. | N=119 (58.3%) | N.D. |
| ≥4 | N=34 (3.3%) |  | N=13 (12.5%) |  | N=85 (41.7%) |  |
| **Median MOG-IgG Titre**<br>**(min-max)** | N.D. | N.D. | N.D. | N.D. | 1280<br>(640-20480) | N.D. |
| **Median AQP4-IgG Titre**<br>**(min-max)** | N.D. | N.D. | 1280<br>(40-20480) | N.D. | N.D. | N.D. |

<sup>a</sup> Healthy controls were selected to match patient sampling patterns; they did not constitute an independent “prediagnostic” cohort, and archived samples were included as required for 1:1 matching.
<sup>b</sup> RIS participants contributed two prospectively collected time points (at RIS diagnosis and at conversion or last follow-up) and were analyzed separately from the archived prediagnostic cohorts.

