## Supplementary Information for "Early immune activation in the prediagnostic phases of immune-mediated neurological diseases"

**Supplementary Tables**

**Table S1: Included Timepoints of MS, NMOSD and MOGAD patients**

| | **Study Cohort** | | | | | |
|---|---|---|---|---|---|---|
| | **MS** | | **NMOSD** | | **MOGAD** | |
| | **MS Patients (N=362)** | **MS Controls (N=362)** | **NMOSD Patients (N=60)** | **NMOSD Controls (N=60)** | **MOGAD Patients (N=145)** | **MOGAD Controls (N=145)** |
| **Time point 1** | | | | | | |
| Included Samples (%) | N=362 (100%) | N=362 (100%) | N=60 (100%) | N=60 (100%) | N=145 (100%) | N=145 (100%) |
| Mean (Min-Max) Months before Disease Diagnosis | 17 (0.04-35.8) | 17 (0.03-46.4) | 22.2 (1.6-75.6) | 22.6 (0.6-78.6) | 8.5 (0.6-36.7) | 8.5 (0.5-35.3) |
| **Time point 2** | | | | | | |
| Included Samples (%) | N=362 (100%) | N=362 (100%) | N=29 (48.3%) | N=29 (48.3%) | N=94 (64.8%) | N=94 (64.8%) |
| Mean (Min-Max) Months before Disease Diagnosis | 48.55 (19.3-90.8) | 46.6 (24-78) | 56.6 (17.1-105.8) | 57.3 (16-107.6) | 62.4 (7.9-120) | 62.4 (7.2-119.7) |
| **Time point 3** | | | | | | |
| Included Samples (%) | N=268 (74%) | N=268 (74%) | N=8 (13.3%) | N=8 (13.3%) | N=12 (8.3%) | N=12 (8.3%) |
| Mean (Min-Max) Months before Disease Diagnosis | 90.1 (55.9-119.2) | 85.8 (45.3-118.5) | 71.9 (34.4-115.5) | 71.3 (36.5-115) | 88.5 (29.4-119.2) | 88.5 (31.1-118.3) |

| | | | | | | |
|---|---|---|---|---|---|---|
| **Time point 4**<br>Included Samples (%)<br>Mean (Min-Max) Months before Disease Diagnosis | N=142 (39.2%)<br>104.1 (97.6-119.7) | N=142 (39.2%)<br>100.8 (65.5-119.9) | N=2 (3.3%)<br>86.8 (85.1-96.4) | N=2 (3.3%)<br>87.3 (84.6-89.8) | N=2 (1.4%)<br>98 (94.4-101.6) | N=2 (1.4%)<br>91.5 (91.2-91.9) |

**Abbreviations:**

**MS:** Multiple Sclerosis**, MOGAD:** Myelin Oligodendrocyte Glycoprotein Antibody–Associated Disease**, NMOSD:** Neuromyelitis Optica Spectrum Disorder.

**Table S2: Characteristics of the Study Cohort**

| | Study Cohort | | | | | |
|---|---|---|---|---|---|---|
| | MS | | NMOSD | | MOGAD | |
| | MS Patients (N=362) | MS Controls (N=362) | NMOSD Patients (N=60) | NMOSD Controls (N=60) | MOGAD Patients (N=145) | MOGAD Controls (N=145) |
| **Median Age (min-max)** | 26.3 (8-67) | 24.9 (12-65) | 49.5 (13-85) | 46.9 (16-83) | 27.3 (4-83) | 26.6 (8-78) |
| **Sex** | | | | | | |
| Female (%) | N=225 (62.2%) | N=225 (62.2%) | N=48 (80%) | N=48 (80%) | N=73 (50.3%) | N=73 (50.3%) |
| Male (%) | N=137 (37.8%) | N=137 (37.8%) | N=12 (20%) | N=12 (20%) | N=72 (49.7%) | N=72 (49.7%) |
| **EBV VCA pos. (%)** | N=362 (100%) | N=362 (100%) | N=52 (86.7%) | N=52 (86.7%) | N=105 (72.4%) | N=105 (72.4%) |
| **EDSS** | | | | | | |
| <4 | N=10 (2.8%) | | N=11 (18.3%) | | N=64 (44.1%) | |
| ≥4 | N=352 (97.2%) | | N=49 (81.7%) | | N=81 (55.9%) | |
| **Median MOG-IgG Titer (min-max)** | N.D. | N.D. | N.D. | N.D. | 1280 (640-20480) | N.D. |
| **Median AQP4-IgG Titer (min-max)** | N.D. | N.D. | 1280 (80-10240) | N.D. | N.D. | N.D. |

**Abbreviations:**

**AQP4:** Aquaporin-4**, EDSS:** expanded disability status scale**, EBV:** Epstein–Barr virus, **MS:** Multiple Sclerosis**, MOG:** Myelin Oligodendrocyte Glycoprotein**, MOGAD:** Myelin Oligodendrocyte Glycoprotein Antibody–Associated Disease**, NMOSD:** Neuromyelitis Optica Spectrum Disorder**, VCA:** viral-capsid antigen.

**Table S3: Peptide Sequences**

| ID | Peptide Sequence |
|---|---|
| **CNS-derived Peptides** | |
| $GlialCAM_{370\text{-}389}$ | ATGRTHSSPPRAPSSPGRSR |
| $CRYAB_{2\text{-}21}$ | DIAIHHPWIRRPFFPFHSPS |
| $MBP_{205\text{-}224}$ | GKGRGLSLSRFSWGAEQQRP |
| $ANO2_{135\text{-}154}$ | PHAGGPGDIELGPLDALEEE |

**Supplementary Figures**

**Figure S1**

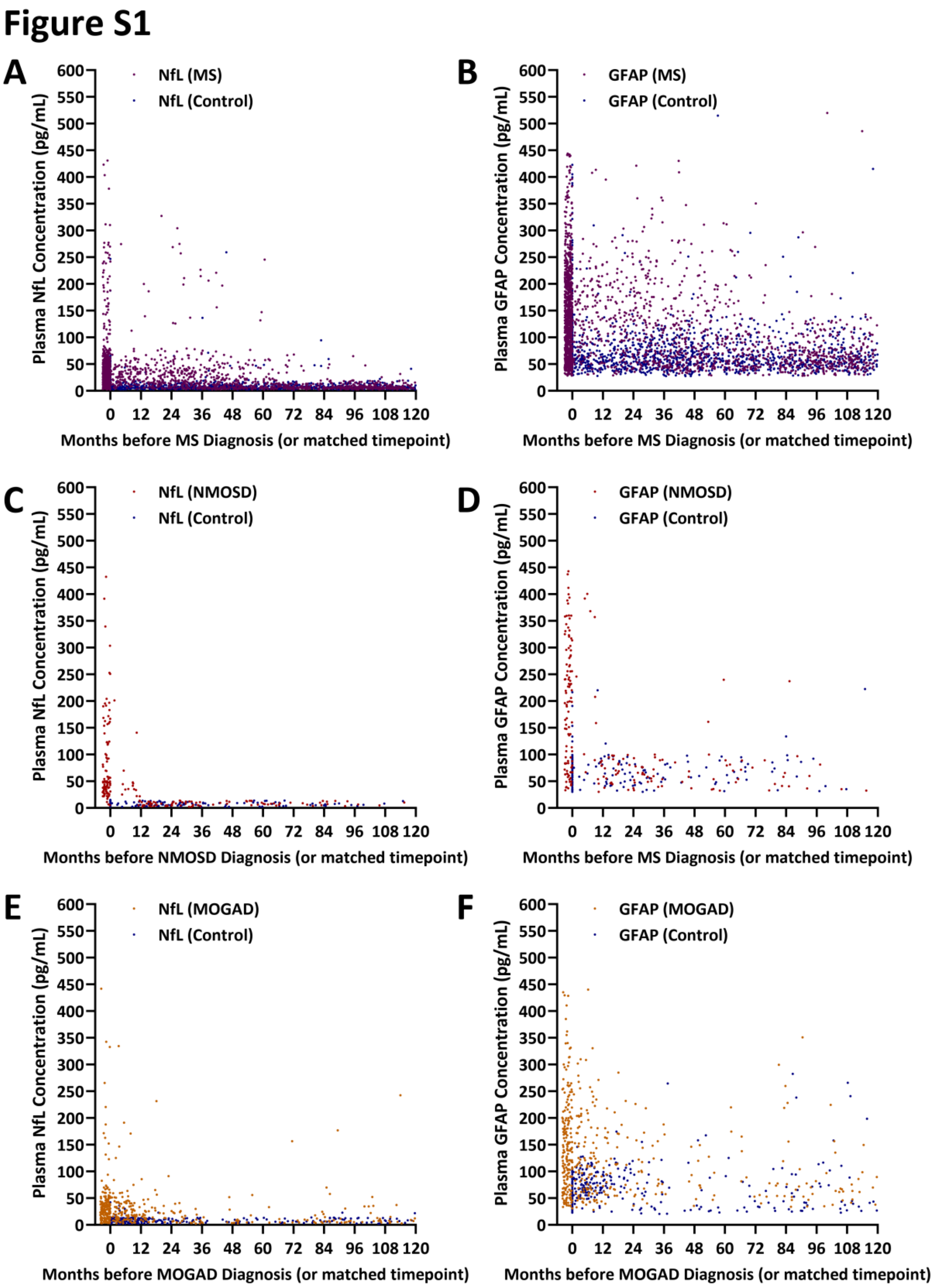

**Figure S1. Temporal dynamics of pNfL and pGFAP in the preclinical phases of MS, NMOSD, and MOGAD.** Plasma samples were collected from (A–B) 1,039 MS patients and matched controls, (C–D) 104 NMOSD patients and matched controls, and (E–F) 204 MOGAD patients and matched controls after the respective disease diagnoses. In addition, retrospective plasma samples were collected from (A–B) 362 MS patients and matched controls (mean 3.1, range 1–4 samples per patient), (C–D) 60 NMOSD patients and matched controls (mean 1.7, range 1–4), and (E–F) 145 MOGAD patients and matched controls (mean 1.7, range 1–4) within ten years before diagnosis or at a matched time point for controls. Concentrations of (A, C, E) pNfL and (B, D, F) pGFAP were analysed in all samples. Each dot represents an individual plasma sample from one future MS, NMOSD, or MOGAD patient or from a matched control. **Abbreviation**: **GFAP**: glial fibrillary acidic protein, **MOGAD:** Myelin Oligodendrocyte Glycoprotein Antibody–Associated Disease, **MS**: Multiple Sclerosis, **NfL**: neurofilament light chain, **NMOSD**: Neuromyelitis Optica Spectrum Disorder.

**Figure S2**

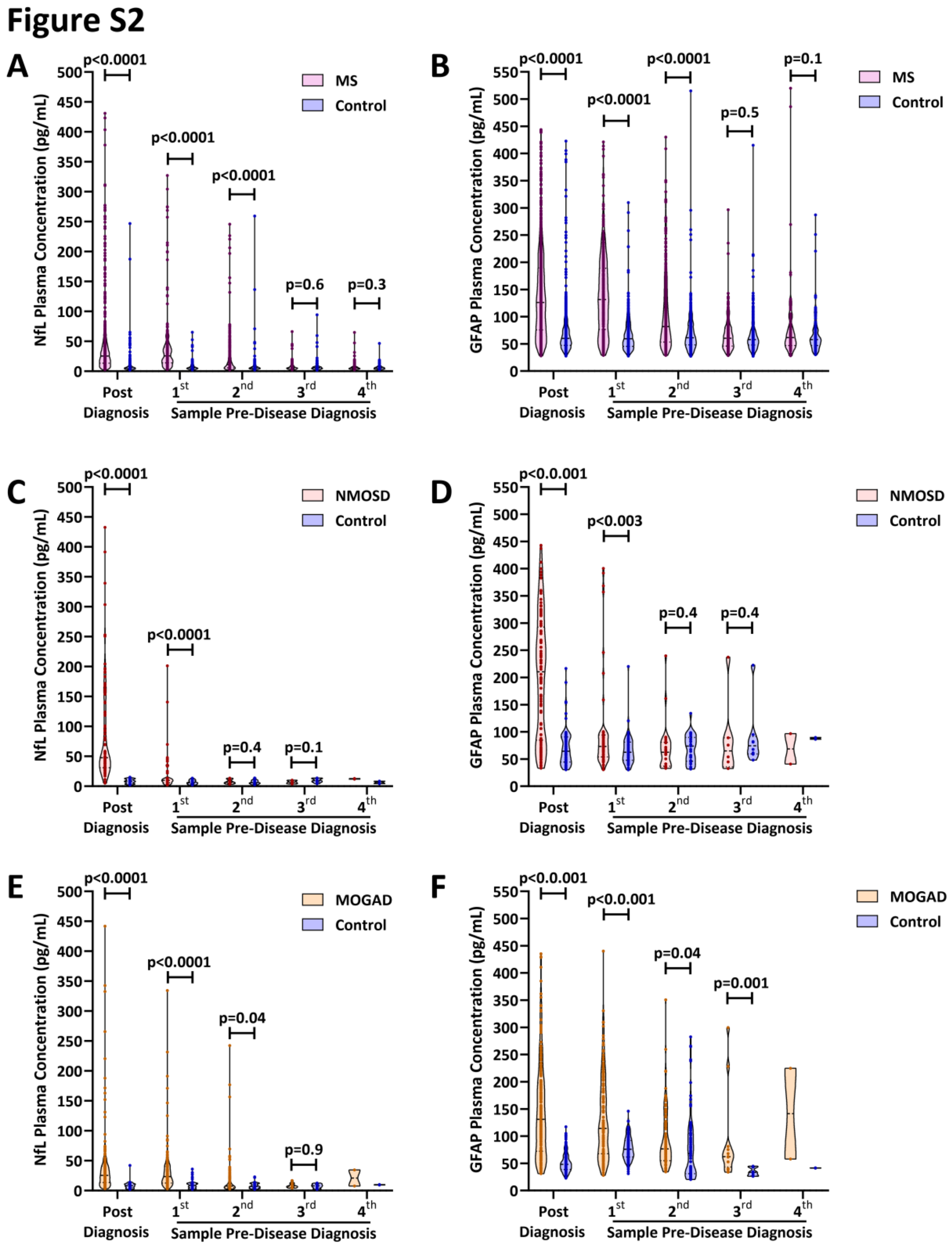

**Figure S2. Temporal dynamics of pNfL and pGFAP in the preclinical phases of MS, NMOSD, and MOGAD.** Plasma samples were collected from (A–B) 1,039 MS patients and matched controls, (C–D) 104 NMOSD patients and matched controls, and (E–F) 204 MOGAD patients and matched controls after the respective disease diagnoses. In addition, between 1–4 retrospective plasma samples were collected from (A–B) 362 MS patients and matched controls (mean 3.1, range 1–4 samples per patient), (C–D) 60 NMOSD patients and matched controls (mean 1.7, range 1–4), and (E–F) 145 MOGAD patients and matched controls (mean 1.7, range 1–4) within ten years before diagnosis or at a matched time point for controls. Violin plots show concentrations of (A, C, E) pNfL and (B, D, F) pGFAP at each assessed time point. Each dot represents an individual plasma sample from a future MS, NMOSD, or MOGAD patient or from a matched control. (A, C, E) pNfL and (B, D, F) pGFAP were compared between patients and matched controls using the Wilcoxon matched-pairs signed-rank test. **Abbreviation**: **GFAP**: glial fibrillary acidic protein, **MOGAD:** Myelin Oligodendrocyte Glycoprotein Antibody–Associated Disease, **MS**: Multiple Sclerosis, **NfL**: neurofilament light chain, **NMOSD**: Neuromyelitis Optica Spectrum Disorder.

**Figure S3**

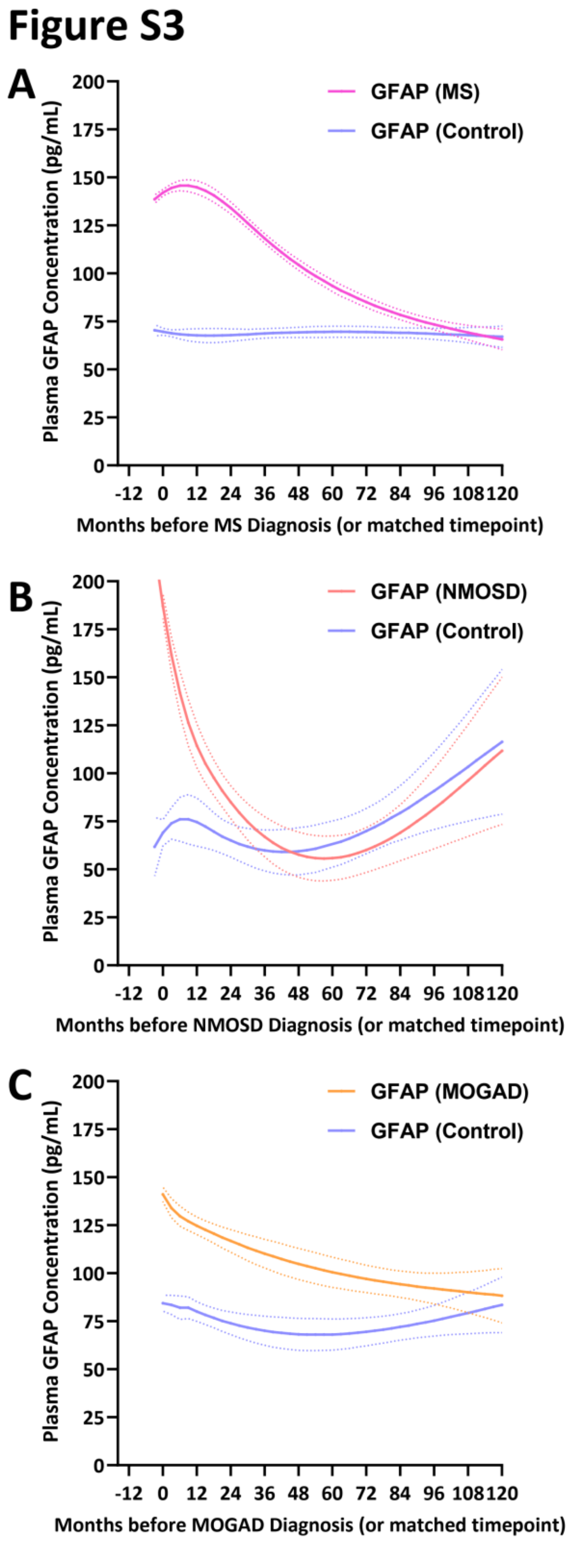

**Figure S3: Temporal dynamics of pGFAP in the preclinical phases of MS, NMOSD, and MOGAD.** Plasma samples were collected from (A) 1,039 MS patients and matched controls, (B) 104 NMOSD patients and matched controls, and (C) 204 MOGAD patients and matched controls after the respective disease diagnoses. In addition, retrospective plasma samples were collected from (A) 362 MS patients and matched controls (mean 3.1, range 1–4 samples per patient), (B) 60 NMOSD patients and matched controls (mean 1.7, range 1–4), and (C) 145 MOGAD patients and matched controls (mean 1.7, range 1–4) within ten years before diagnosis or at a matched time point for controls. Concentrations of pGFAP were analysed in all samples. pGFAP concentrations over time were modelled using a linear mixed model to account for repeated sampling. Means and standard errors are shown. **Abbreviations**: **GFAP**: glial fibrillary acidic protein, **MOGAD:** Myelin Oligodendrocyte Glycoprotein Antibody–Associated Disease, **MS**: Multiple Sclerosis, **NMOSD**: Neuromyelitis Optica Spectrum Disorder.

**Figure S4**

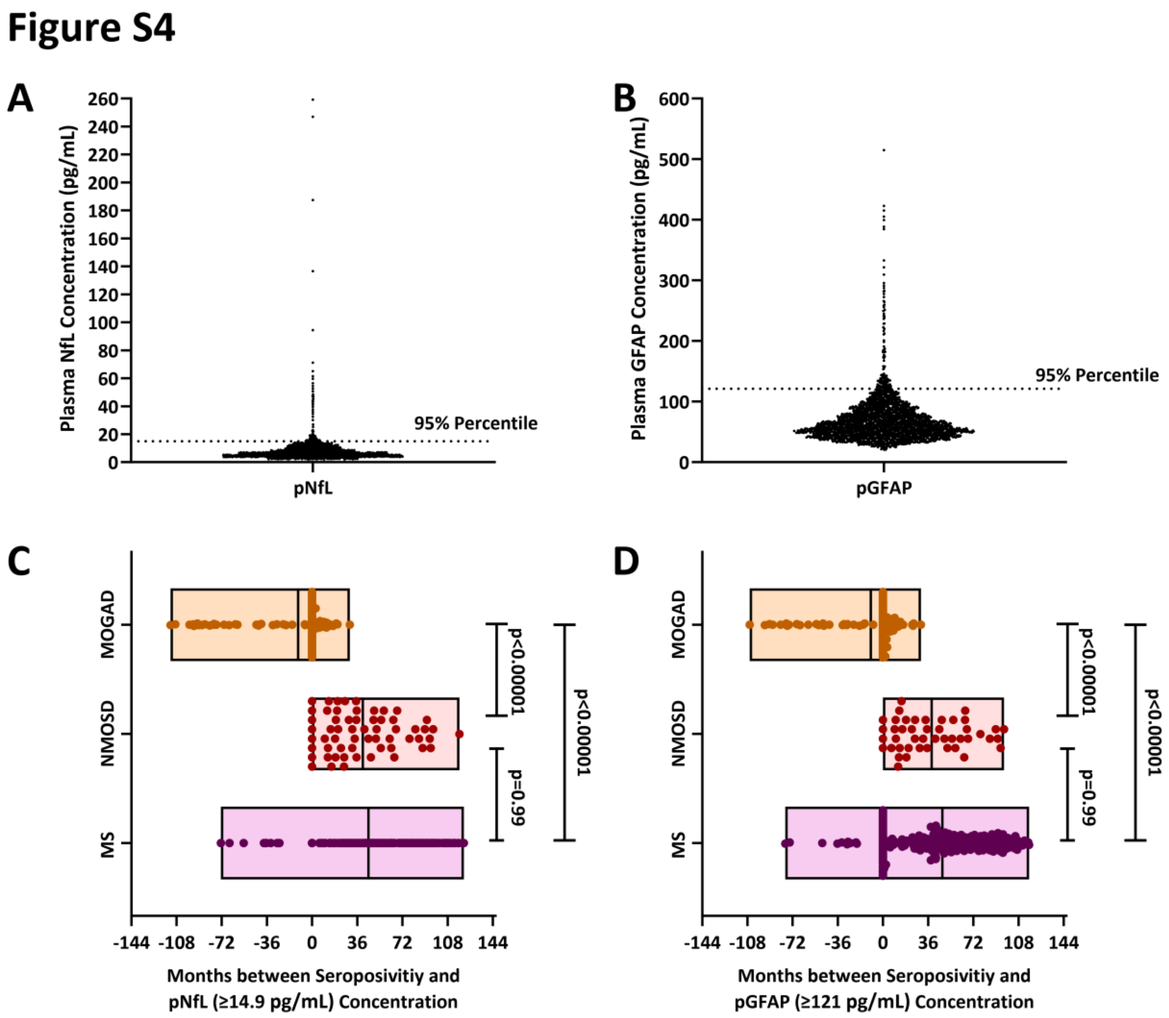

**Figure S4: High-level pNfL and pGFAP emerge after EBNA-1 and AQP4 antibodies in MS and NMOSD patients.** (A–B) Definition of high-level (A) pNfL and (B) pGFAP concentrations. pNfL and pGFAP were measured in 2,831 plasma samples from 1,288 healthy controls. High-level pNfL (≥14.91 pg/mL) and pGFAP (≥121 pg/mL) concentrations were defined as values exceeding those observed in 95% of plasma samples from healthy controls. (C–D) High-level pNfL and pGFAP concentrations emerge after EBNA-1–IgG and AQP4-IgG in MS and NMOSD patients. Plasma samples were collected from 1,039 MS patients, 104 NMOSD patients, and 204 MOGAD patients after the respective disease diagnoses. In addition, retrospective plasma samples were collected from 362 MS patients (mean 3.1, range 1–4 samples per patient), 60 NMOSD patients (mean 1.7, range 1–4), and 145 MOGAD patients (mean 1.7, range 1–4) within ten years before diagnosis. pNfL and pGFAP were assessed in all plasma samples. All plasma samples from future MS, NMOSD, and MOGAD patients were tested for the presence of EBNA-1–IgG, AQP4-IgG, and MOG-IgG, respectively. Floating bars show the mean time (min–max) between the first detection of EBNA-1–IgG, AQP4-IgG, and MOG-IgG in MS, NMOSD, and MOGAD, as well as the first detection of pNfL ≥14.91 pg/mL or pGFAP ≥121 pg/mL. Each dot represents an individual patient. Data were analyzed using the Kruskal–Wallis test followed by Dunn's multiple comparisons test. **Abbreviations**: **AQP4**: Aquaporin-4, **EBNA-1**: Epstein-Barr Virus Nuclear Antigen 1, **GFAP**: glial fibrillary acidic protein**, MOG**: Myelin Oligodendrocyte Glycoprotein, **MOGAD**: Myelin Oligodendrocyte Glycoprotein Antibody–Associated Disease, **MS**: Multiple Sclerosis, **NfL**: neurofilament light chain, **NMOSD**: Neuromyelitis Optica Spectrum Disorder..

**Figure S5**

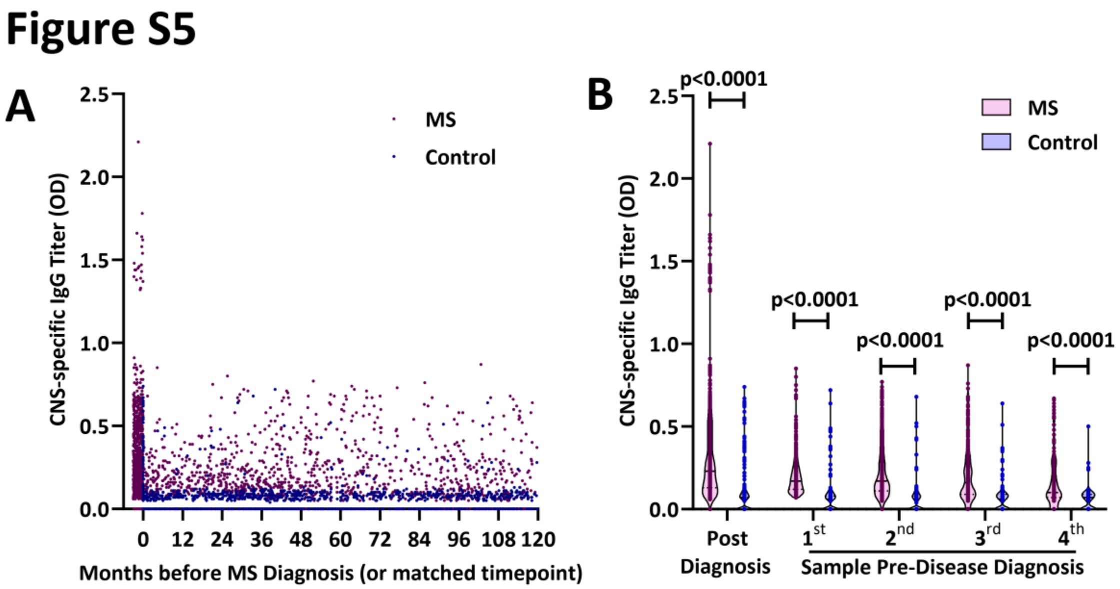

**Figure S5**: **Temporal dynamics of CNS-specific IgG antibodies in the preclinical phase of MS.** Plasma samples were collected from 1,039 MS patients and matched controls. In addition, retrospective plasma samples were collected from 362 MS patients and matched controls (mean 3.1, range 1–4 samples per patient) within ten years before diagnosis or at a matched time point for controls. Plasma CNS-specific IgG antibody titers were analysed in all samples. (A) Each dot represents the CNS-specific IgG titer in an individual plasma sample from a future MS patient or a matched control. (B) Violin plots show CNS-specific IgG antibody titers at each assessed time point. CNS-specific IgG antibody titers were compared between patients and matched controls using the Wilcoxon matched-pairs signed-rank test. **Abbreviation**: **CNS:** central nervous system, **MS**: Multiple Sclerosis. **OD**: Optical Density.

**Figure S6**

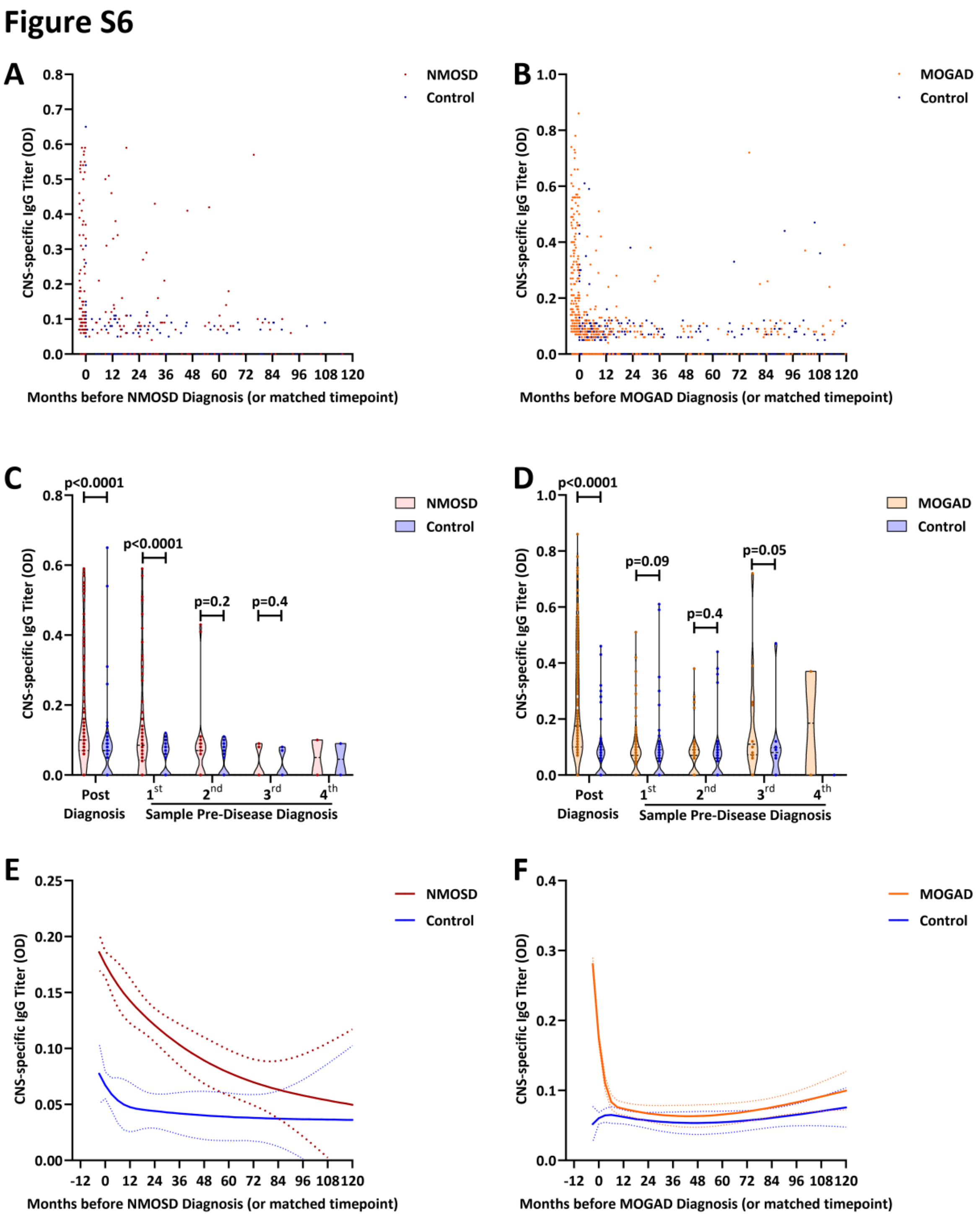

**Figure S6. Temporal dynamics of CNS-specific IgG antibodies in the preclinical phases of NMOSD and MOGAD.** Plasma samples were collected from (A, C, E) 104 NMOSD patients and matched controls and from (B, D, F) 204 MOGAD patients and matched controls after the respective disease diagnoses. In addition, retrospective plasma samples were collected from (A,

C, E) 60 NMOSD patients and matched controls (mean 1.7, range 1–4 samples per patient) and from (B, D, F) 145 MOGAD patients and matched controls (mean 1.7, range 1–4) within ten years before diagnosis or at a matched time point for controls. (A–B) Each dot represents an individual plasma sample from a future (A) NMOSD or (B) MOGAD patient or from a matched control. (C–D) Violin plots show CNS-specific IgG antibody titers at each assessed time point in future (C) NMOSD or (D) MOGAD patients. CNS-specific IgG titers were compared between patients and matched controls using the Wilcoxon matched-pairs signed-rank test. (E–F) CNS-specific IgG antibody titers in future (E) NMOSD or (F) MOGAD patients over time were analyzed using a linear mixed model to account for repeated sampling. **Abbreviations**: **CNS:** central nervous system, **MOGAD:** Myelin Oligodendrocyte Glycoprotein Antibody–Associated Disease, **NMOSD**: Neuromyelitis Optica Spectrum Disorder, **OD:** Optical Density.

**Figure S7**

**Figure S7**

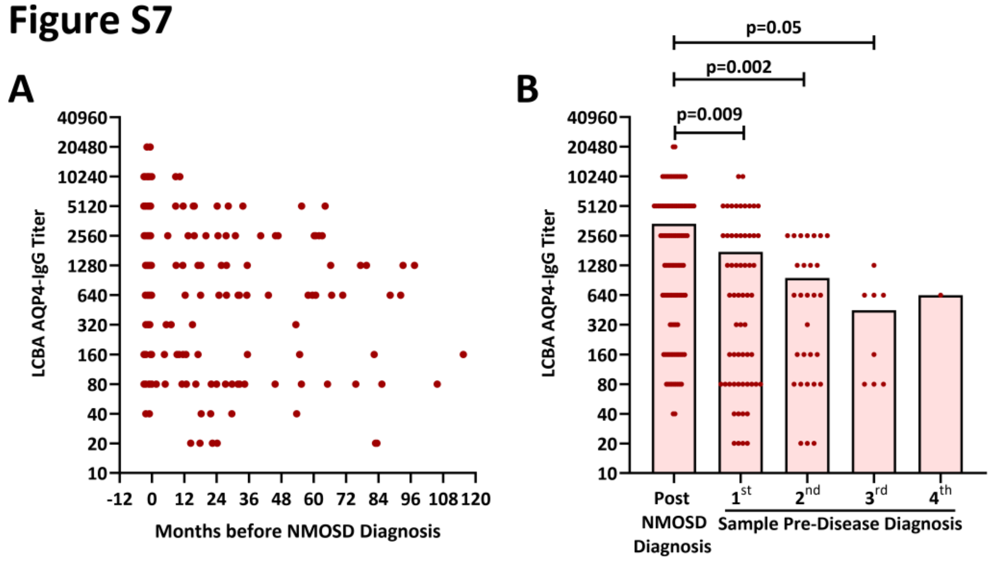

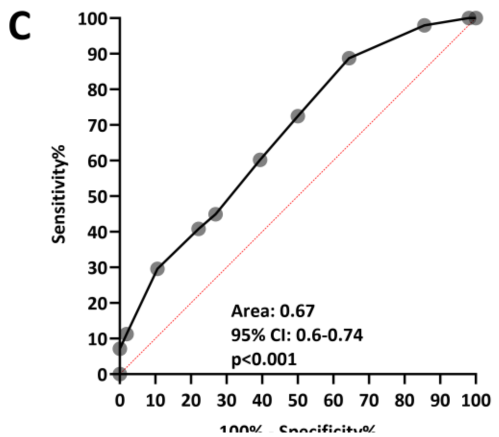

| | Sensitivity% | 95% CI | Specificity% | 95% CI |
|---|---|---|---|---|
| < 30.00 | 7.1 | 3.503% to 14.02% | 100.0 | 96.44% to 100.0% |
| < 60.00 | 11.2 | 6.384% to 18.99% | 98.1 | 93.26% to 99.66% |
| < 120.0 | 29.6 | 21.46% to 39.26% | 89.4 | 82.05% to 93.99% |
| < 240.0 | 40.8 | 31.61% to 50.71% | 77.9 | 69.00% to 84.79% |
| < 480.0 | 44.9 | 35.43% to 54.75% | 73.1 | 63.84% to 80.67% |
| < 960.0 | 60.2 | 50.30% to 69.33% | 60.6 | 50.97% to 69.43% |
| < 1920 | 72.5 | 62.88% to 80.32% | 50.0 | 40.56% to 59.44% |
| < 3840 | 88.8 | 81.01% to 93.62% | 35.6 | 27.04% to 45.14% |
| < 7680 | 98.0 | 92.86% to 99.64% | 14.4 | 8.940% to 22.44% |
| < 15360 | 100.0 | 96.23% to 100.0% | 1.9 | 0.3417% to 6.742% |

**Figure S7: Temporal dynamics of AQP4--IgG in the preclinical phase of NMOSD.** (A) AQP4-specific IgG titers were assessed and quantified in 104 NMOSD patients. In addition, AQP4-specific IgG titers were measured in retrospective plasma samples from 60 NMOSD patients and matched controls (mean 1.7, range 1–4 samples per patient) within ten years before diagnosis or at a matched time point for controls. Each dot represents the AQP4-specific IgG titer in an individual plasma sample from a future NMOSD patient. (B) Box plots represent the mean AQP4-specific IgG titer at each assessed time point, with individual plasma samples shown as dots. Titers at each time point were compared using the Mann–Whitney test. (C) ROC analysis discriminating AQP4-specific IgG titers before (n = 60 patients) and after (n = 104 patients) NMOSD diagnosis. **Abbreviations**: **AQP4**: Aquaporin-4, **LCBA**: live cell-based assay, **NMOSD**: Neuromyelitis Optica Spectrum Disorder, **ROC**: Receiver operating characteristic.

**Figure S8**

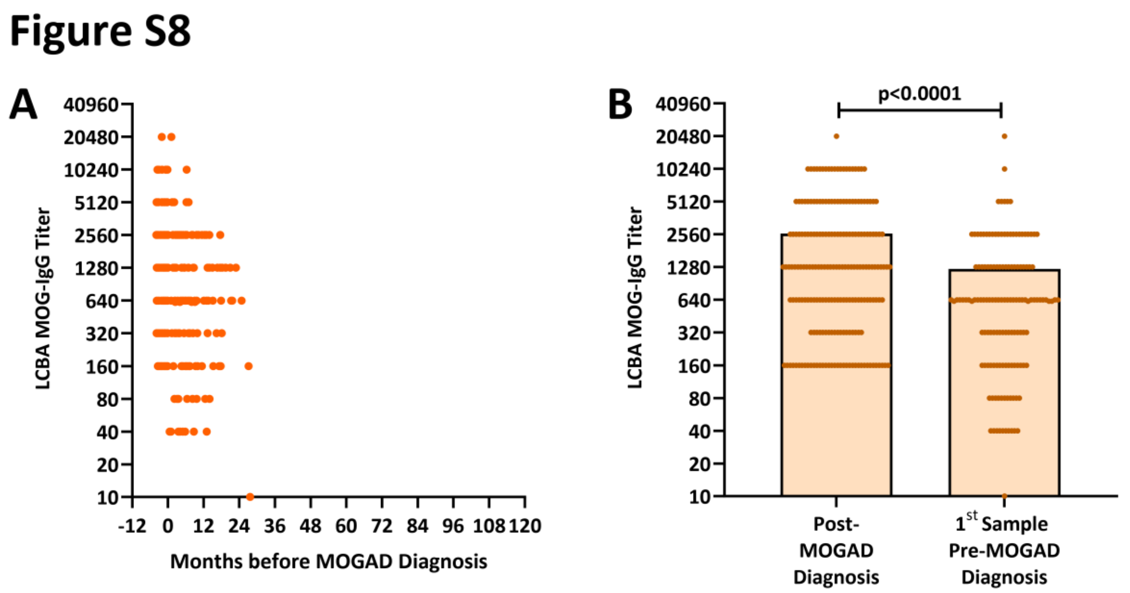

**Figure S8: Temporal dynamics of MOG-IgG in the preclinical phase of MOGAD.** (A) MOG-specific IgG titers were assessed and quantified in 204 MOGAD patients. In addition, MOG-specific IgG titers were measured in retrospective plasma samples from 145 MOGAD patients and matched controls (mean 1.7, range 1–4 samples per patient) within ten years before diagnosis or at a matched time point for controls. Each dot represents the MOG-specific IgG titer in an individual plasma sample from a future MOGAD patient. (B) Box plots represent the mean MOG-specific IgG titer at each assessed time point, with individual plasma samples shown as dots. Titers at each time point were compared using the Mann–Whitney test. **Abbreviations**: **LCBA**: live cell-based assay, **MOG**: Myelin Oligodendrocyte Glycoprotein, **MOGAD**: Myelin Oligodendrocyte Glycoprotein Antibody–Associated Disease, **NMOSD**: Neuromyelitis Optica Spectrum Disorder.
